# Fundamental principles of epidemic spread highlight the immediate need for large-scale serological surveys to assess the stage of the SARS-CoV-2 epidemic

**DOI:** 10.1101/2020.03.24.20042291

**Authors:** José Lourenço, Robert Paton, Craig Thompson, Paul Klenerman, Sunetra Gupta

## Abstract

The spread of a novel pathogenic infectious agent eliciting protective immunity is typically characterised by three distinct phases: (I) an initial phase of slow accumulation of new infections (often undetectable), (II) a second phase of rapid growth in cases of infection, disease and death, and (III) an eventual slow down of transmission due to the depletion of susceptible individuals, typically leading to the termination of the (first) epidemic wave. Before the implementation of control measures (e.g. social distancing, travel bans, etc) and under the assumption that infection elicits protective immunity, epidemiological theory indicates that the ongoing epidemic of SARS-CoV-2 will conform to this pattern.

Here, we calibrate a susceptible-infected-recovered (SIR) model to data on cumulative reported SARS-CoV-2 associated deaths from the United Kingdom (UK) and Italy under the assumption that such deaths are well reported events that occur only in a vulnerable fraction of the population. We focus on model solutions which take into consideration previous estimates of critical epidemiological parameters such as the basic reproduction number (R_0_), probability of death in the vulnerable fraction of the population, infectious period and time from infection to death, with the intention of exploring the sensitivity of the system to the actual fraction of the population vulnerable to severe disease and death.

Our simulations are in agreement with other studies that the current epidemic wave in the UK and Italy in the absence of interventions should have an approximate duration of 2-3 months, with numbers of deaths lagging behind in time relative to overall infections. Importantly, the results we present here suggest the ongoing epidemics in the UK and Italy started at least a month before the first reported death and have already led to the accumulation of significant levels of herd immunity in both countries. There is an inverse relationship between the proportion currently immune and the fraction of the population vulnerable to severe disease.

This relationship can be used to determine how many people will require hospitalisation (and possibly die) in the coming weeks if we are able to accurately determine current levels of herd immunity. There is thus an urgent need for investment in technologies such as virus (or viral pseudotype) neutralization assays and other robust assays which provide reliable read-outs of protective immunity, and for the provision of open access to valuable data sources such as blood banks and paired samples of acute and convalescent sera from confirmed cases of SARS-CoV-2 to validate these. Urgent development and assessment of such tests should be followed by rapid implementation at scale to provide real-time data. These data will be critical to the proper assessment of the effects of social distancing and other measures currently being adopted to slow down the case incidence and for informing future policy direction.

**Disclaimer:** (a) This material is not final and is subject to be updated any time. (b) Code used will be made available as soon as possible. (c) Contact for press enquiries: Cairbre Sugrue, +44 (0)7502 203 769.

## Main Text

Our overall approach rests on the assumption that only a very small proportion of the population is at risk of hospitalisable illness. This proportion is itself only a fraction of the risk groups already well described in the literature [1–4], including the elderly and those carrying critical comorbidities (e.g. asthma). We used a susceptible-infectious-recovered framework (SIRf) to examine the effects of varying the vulnerable fraction of the population on the transmission dynamics of SARS-CoV-2, fixing other model parameters to ranges supported by previous studies **(Table 1)**. We fit the model to cumulative deaths in the United Kingdom and Italy in the first 15 days following the first recorded death to avoid any potential effects of local control strategies implemented since that time.

**Table 1.** Model variables and parameters. M=mean. SD=standard deviation. S=scale. R=rate.

| Variable / Parameter |  | Assumptions / Priors | Support |
| --- | --- | --- | --- |
| proportion infectious | $y$ | equation 1 | --- |
| proportion of population no longer susceptible | $z$ | equation 2 | --- |
| cumulative deaths | $\Lambda$ | equation 3 | --- |
| time (day) of introduction | $\tau$ | Uniform distribution $(-\infty, +\infty)$ | --- |
| basic reproduction number | $R_0$ | Gaussian distributions:<br>G1(M=2.25, SD=0.025),<br>G2(M=2.75, SD=0.025) | [11–13] |
| infectious period (days) | $1/\sigma$ | Gaussian distribution<br>G(M=4.5, SD=1) | [11,14–16] |
| transmission coefficient | $\beta$ | $\beta = \sigma R_0$ | --- |
| time (days) between infection and death | $\psi$ | Gaussian distribution<br>G(M=17, SD=2) | [14] |
| probability of dying with severe disease | $\theta$ | Gaussian distribution<br>G(M=0.14, SD=0.007) | [1,2,11,17] |
| proportion of population at risk of severe disease | $\rho$ | Gamma distribution<br>G1(S=5, R=5/0.1),<br>G2(S=5, R=5/0.03),<br>G3(S=5, R=5/0.01),<br>G4(S=5, R=5/0.001) | --- |
| population size | $N$ | UK 66.87M, Italy 60M | --- |

Three different scenarios under which the model closely reproduces the reported death counts in the UK up to 19/03/2020 are presented in **Figure 1**. Red and green colours represent solutions attached respectively to transmission scenarios with R_0_=2.75 and R_0_=2.25 (reflecting variation in estimates of R_0_ in literature) with the proportion of the population at risk being distributed around 1%. The model output (posterior) for time of introduction (the start of transmission) place this event a couple of days after the first confirmed case in the country, and over a month before the first confirmed death **(Figures 1E-F)**. In both R_0_ scenarios, by the time the first death was reported (05/03/2020), thousands of individuals (∼0.08%) would have already been infected with the virus (as also suggested by [5]). By 19/03/2020, approximately 36% (R_0_=2.25) and 40% (R_0_=2.75) of the population would have already been exposed to SARS-CoV-2. Running the same model with R_0_=2.25 and the proportion of the population at risk of severe disease being distributed around 0.1%, places the start of transmission at 4 days prior to first case detection and 38 days before the first confirmed death and suggests that 68% would have been infected by 19/03/2020.

**Figure 1.**
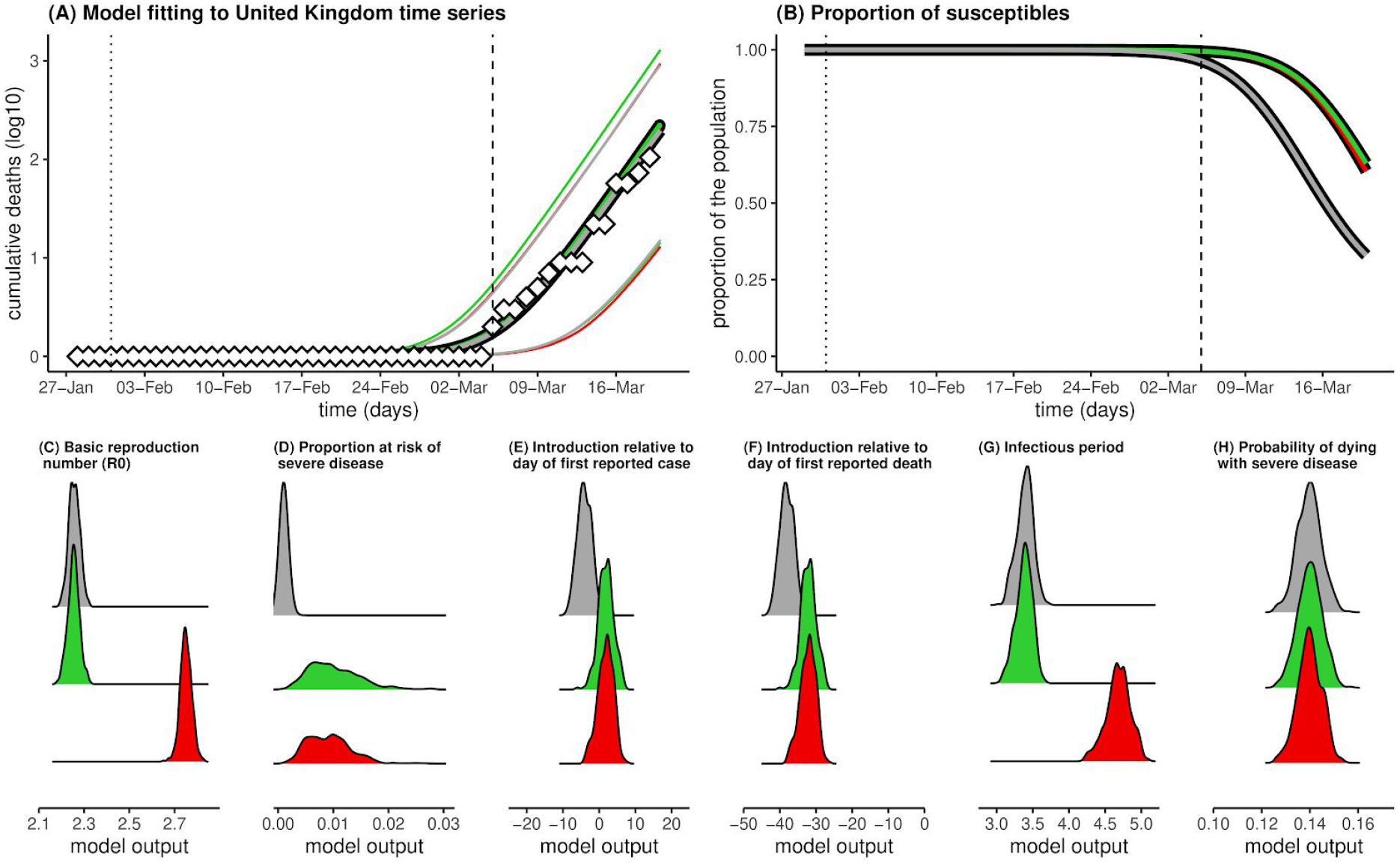
Results for the United Kingdom for three scenarios: R_0_ = 2.25 and ρ = 0.001 (grey), R_0_ = 2.25 and ρ = 0.01 (green), and R_0_ = 2.75 and ρ = 0.01 (red). MCMC ran for 1 million steps. Results presented are the posteriors (model output) using 1000 samples after a burnout of 50% **(A)** Model fits showing reported (diamonds) and model (lines) cumulative death counts. Deaths are log10 transformed for visualisation. **(B)** Mean proportion of the population still susceptible to infection (1-z, see Model). **(A-B)** Vertical lines mark the date of the first confirmed case (dotted) and date of first confirmed death (dashed). **(C)** Posteriors for R0, (**D**) proportion of population at risk of severe disease (ρ), (**E**) Time of introduction relative to the date of the first reported case, (**F)** Time of introduction relative to the date of first reported death, (**G**) infectious period, (**H**) probability of dying with severe disease.

The results of the same exercise for Italy (**Figure 2)** place the time of introduction around 10 days before the first confirmed case, and around a month before the first confirmed death **(Figures 2E-F)** when the proportion of the population at risk of severe disease is around 1%. By 06/03/2020, approximately 45 days post introduction, the model suggests that approximately 60% (R_0_ = 2.25) and 64% (R_0_ = 2.75) of the population would have already been exposed to SARS-CoV-2. When the proportion of the population at risk is around 0.1%, the start of transmission is likely to have occurred 17 days prior to first case detection and 38 days before the first confirmed death with 80% already infected by 06/03/2020.

**Figure 2.**
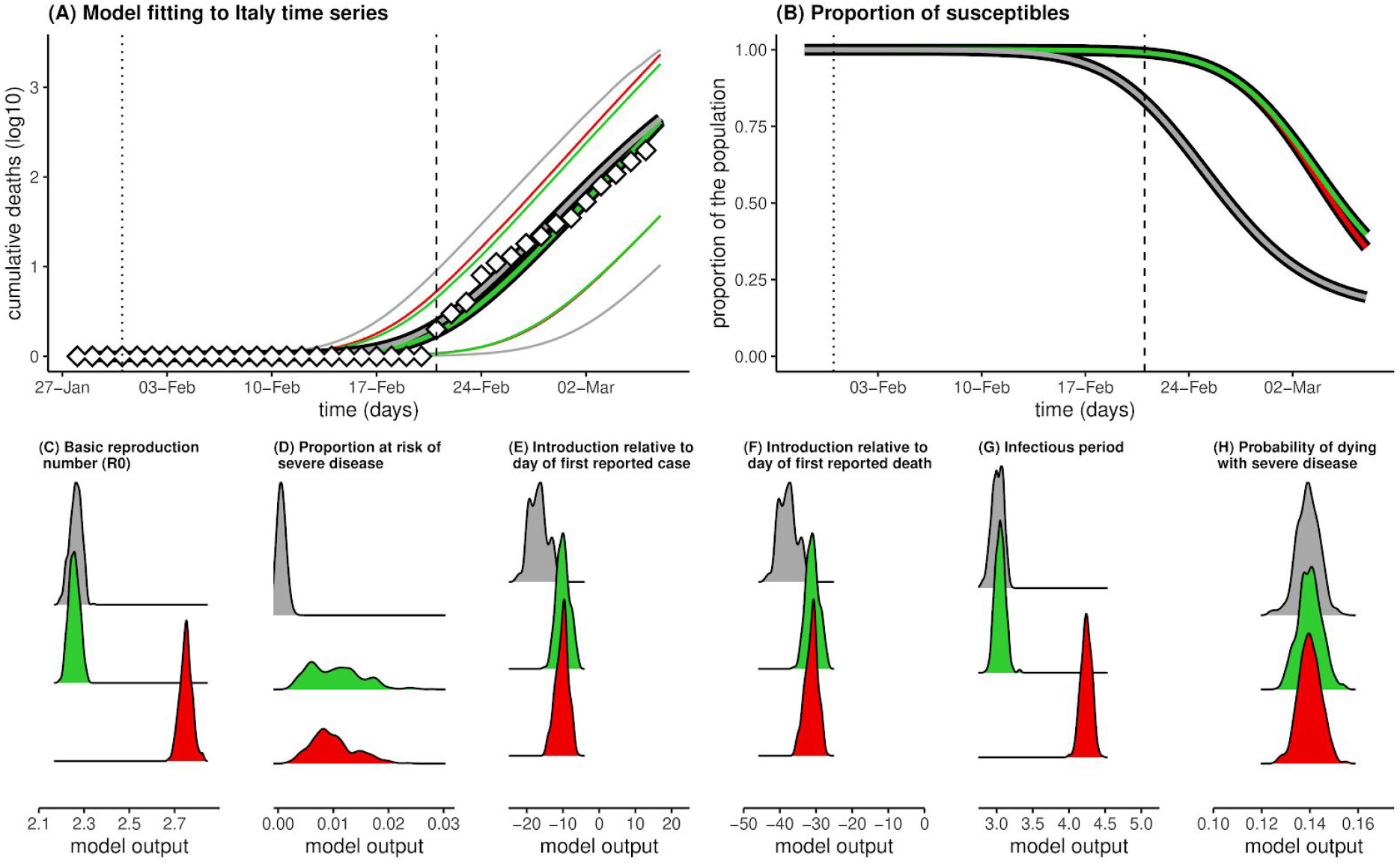
Results for Italy.(A-H) Legend as in Figure 1.

Overall, these results underscore the dependence of the inferred epidemic curve on the assumed fraction of the population vulnerable to severe disease (ρ) showing significant population level immunity accruing by mid March in the UK as ρ is decreased to plausible values (**Figure 3**). They also suggest a way of determining this fraction by measuring the proportion of the population already exposed to SARS-CoV-2.

**Figure 3.**
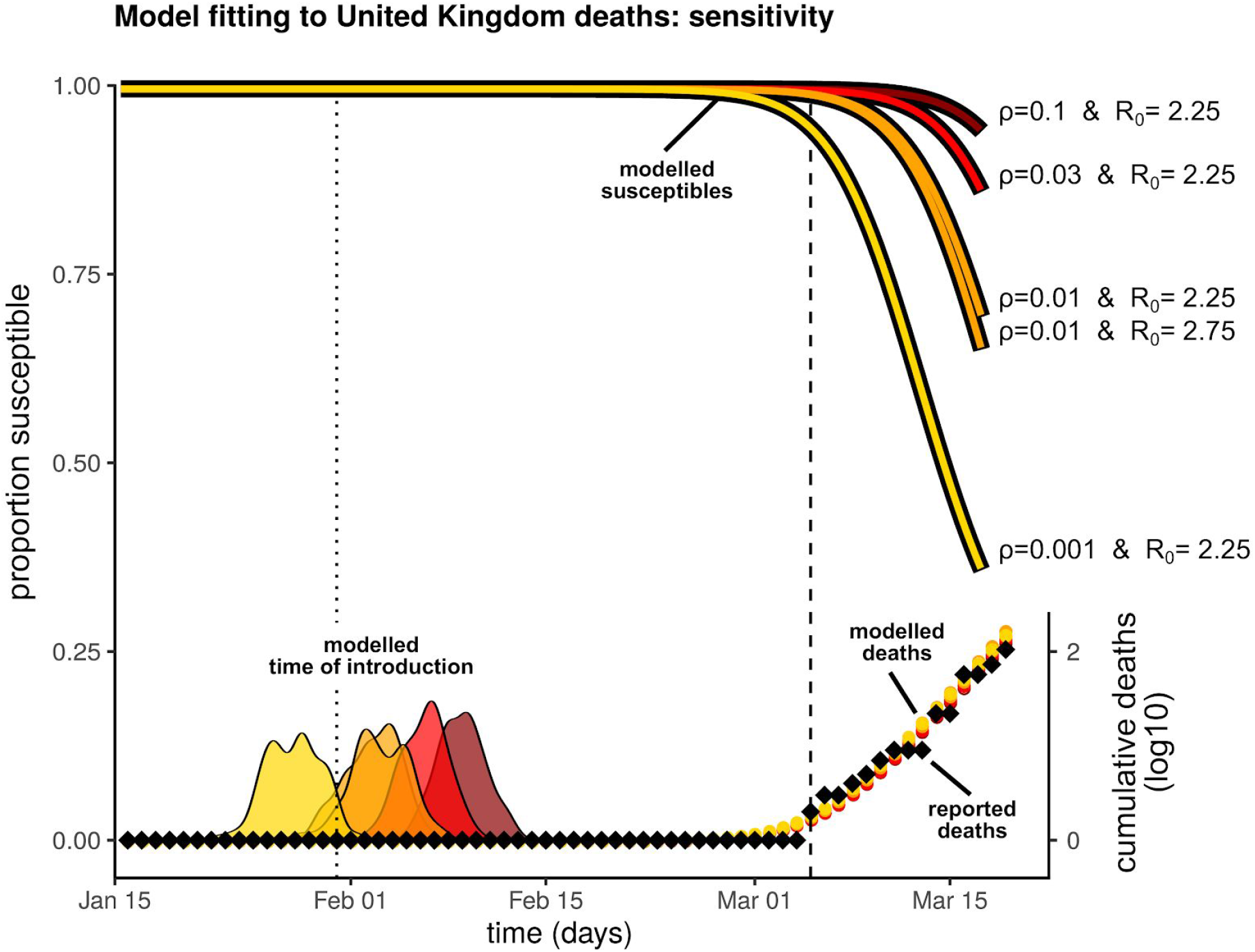
Sensitivity of results to the fraction of the population vulnerable to severe disease. Four scenarios are presented: R_0_ = 2.25 and ρ = 0.1 (dark red), R_0_ = 2.25 and ρ = 0.03 (red), R_0_ ∈ {2.25, 2.75} and ρ = 0.01 (both orange), R_0_ = 2.25 and ρ = 0.001 (yellow). MCMC ran for 1 million steps. Posteriors (model output) were obtained using 1000 samples after a burnout of 50%. Vertical lines mark the date of the first confirmed case (dotted) and date of first confirmed death (dashed).

## Model

We model a susceptible-infectious-recovered framework (SIRf) [6] to simulate the spread of SARS-CoV-2. The population is separated into those currently contributing to transmission (y, **equation 1**) and those not available for infection (z, **equation 2**). Cumulative death counts (Λ, **equation 3**) are obtained by considering that mortality occurs with probability θ, on a proportion of the population that is at risk of severe disease (ρ) among those already exposed (z); we consider the delay between the time of infection and of death (ψ) as a combination of incubation period and time to death after onset of symptoms. The small proportion of the population that is at risk of severe disease (ρ) is an aggregate model parameter, taking into consideration both a potentially lower risk of infection than the rest of the population, as well as the actual risk of severe disease.

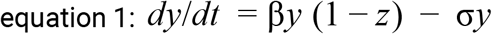

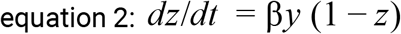

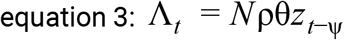

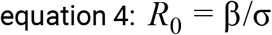

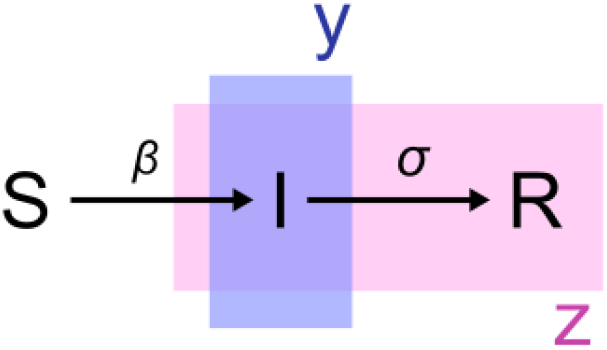

Model output on cumulative death counts (Λ) is fitted to the reported time series of deaths (see Data) using a Bayesian MCMC approach previously implemented in other modelling studies [7–10]. Model variables are summarized in **Table 1**.

## Data

### cumulative number of deaths

#### Italy

A time series was obtained from the Italian Department of Civil Protection GitHub repository [18] (accessed on 17/03/2020). We trimmed the data to the first 15 days of death counts above zero (21/02/2020 to 06/03/2020) to include only the initial increase free of effects from local control measures.

#### UK

A time series was obtained from the John Hopkins University Centre for Systems Science and Engineering COVID-19 GitHub repository [19](accessed 19/03/2020). We trimmed the data to the first 15 days of death counts above zero (05/03/2020 to 19/03/2020) to include only the initial increase free of effects from local control measures.

## Data Availability

The data included in this draft manuscript is from open-access data repositories.

## References

1. Report of the WHO-China Joint Mission on Coronavirus Disease 2019 (COVID-19). 16-24 February 2020. Available: https://www.who.int/docs/default-source/coronaviruse/who-china-joint-mission-on-covid-19-final-report.pdf

2. Verity R, Okell LC, Dorigatti I, Winskill P, Whittaker C, Imai N, et al. Estimates of the severity of COVID-19 disease. medRxiv. 2020. doi:10.1101/2020.03.09.20033357

3. Yang J, Zheng Y, Gou X, Pu K, Chen Z, Guo Q, et al. Prevalence of comorbidities in the novel Wuhan coronavirus (COVID-19) infection: a systematic review and meta-analysis. Int J Infect Dis. 2020. doi:10.1016/j.ijid.2020.03.017

4. Zhou F, Yu T, Du R, Fan G, Liu Y, Liu Z, et al. Clinical course and risk factors for mortality of adult inpatients with COVID-19 in Wuhan, China: a retrospective cohort study. The Lancet. 2020. doi:10.1016/s0140-6736(20)30566-3

5. Jombart T, van Zandvoort K, Russell T, Jarvis C, Gimma A, Abbott S, et al. Inferring the number of COVID-19 cases from recently reported deaths. doi:10.1101/2020.03.10.20033761

6. Anderson RM, Anderson B, May RM. Infectious Diseases of Humans: Dynamics and Control. Oxford University Press; 1992.

7. Lourenço J, Obolski U, Swarthout TD, Gori A, Bar-Zeev N, Everett D, et al. Determinants of high residual post-PCV13 pneumococcal vaccine-type carriage in Blantyre, Malawi: a modelling study. BMC Med. 2019;17: 219.

8. McNaughton AL, Lourenço J, Hattingh L, Adland E, Daniels S, Van Zyl A, et al. HBV vaccination and PMTCT as elimination tools in the presence of HIV: insights from a clinical cohort and dynamic model. BMC Med. 2019;17: 43.

9. Lourenço J, de Lima MM, Faria NR, Walker A, Kraemer MUG, Villabona-Arenas CJ, et al. Epidemiological and ecological determinants of Zika virus transmission in an urban setting. eLife. 2017. doi:10.7554/elife.29820

10. Faria NR, da Costa AC, Lourenço J, Loureiro P, Lopes ME, Ribeiro R, et al. Genomic and epidemiological characterisation of a dengue virus outbreak among blood donors in Brazil. Sci Rep. 2017;7: 15216.

11. Kucharski AJ, Russell TW, Diamond C, Liu Y, Edmunds J, Funk S, et al. Early dynamics of transmission and control of COVID-19: a mathematical modelling study. Lancet Infect Dis. 2020. doi:10.1016/S1473-3099(20)30144-4

12. Park SW, Cornforth DM, Dushoff J, Weitz JS. The time scale of asymptomatic transmission affects estimates of epidemic potential in the COVID-19 outbreak. medRxiv. 2020. doi:10.1101/2020.03.09.20033514

13. Hellewell J, Abbott S, Gimma A, Bosse NI, Jarvis CI, Russell TW, et al. Feasibility of controlling COVID-19 outbreaks by isolation of cases and contacts. Lancet Glob Health. 2020;8: e488–e496.

14. Linton NM, Kobayashi T, Yang Y, Hayashi K, Akhmetzhanov AR, Jung S-M, et al. Incubation Period and Other Epidemiological Characteristics of 2019 Novel Coronavirus Infections with Right Truncation: A Statistical Analysis of Publicly Available Case Data. J Clin Med Res. 2020;9. doi:10.3390/jcm9020538

15. Li Q, Guan X, Wu P, Wang X, Zhou L, Tong Y, et al. Early Transmission Dynamics in Wuhan, China, of Novel Coronavirus-Infected Pneumonia. N Engl J Med. 2020. doi:10.1056/NEJMoa2001316

16. Woelfel R, Corman VM, Guggemos W, Seilmaier M, Zange S, Mueller MA, et al. Clinical presentation and virological assessment of hospitalized cases of coronavirus disease 2019 in a travel-associated transmission cluster. doi:10.1101/2020.03.05.20030502

17. Russell TW, Hellewell J, Jarvis CI, van-Z Voort K, Abbott S, et al. Estimating the infection and case fatality ratio for COVID-19 using age-adjusted data from the outbreak on the Diamond Princess cruise ship. medRxiv. 2020. doi:10.1101/2020.03.05.20031773

18. COVID-19 Italia - Monitoraggio situazione; Presidenza del Consiglio dei Ministri - Dipartimento della Protezione Civile. Available: https://github.com/pcm-dpc/COVID-19

19. Novel Coronavirus (COVID-19) Cases, provided by CSSE at Johns Hopkins University. Available: https://github.com/CSSEGISandData/COVID-19

